# Systematic analysis of short tandem repeats in 38,095 exomes provides an additional diagnostic yield

**DOI:** 10.1101/2020.10.19.20211144

**Authors:** Bart P.G.H. van der Sanden, Jordi Corominas, Michelle de Groot, Maartje Pennings, Rowdy P.P. Meijer, Nienke Verbeek, Bart van de Warrenburg, Meyke Schouten, Helger G. Yntema, Lisenka E.L.M. Vissers, Erik-Jan Kamsteeg, Christian Gilissen

## Abstract

**Purpose:** The expansion of specific short tandem repeats (STRs) can lead to approximately 30 different human genetic disorders. Despite extensive application of exome sequencing (ES) in routine diagnostic genetic testing, STRs are not routinely identified from these data.

**Methods:** We assessed diagnostic utility by applying ExpansionHunter to 2,867 exomes from movement disorder patients and 35,228 other clinical exomes.

**Results:** We identified 36 movement disorder patients with a possible aberrant STR length. Validation by PCR and/or repeat-primed PCR technologies confirmed the presence of aberrant expansion alleles for 11 (31%). For seven of these patients the genotype was compatible with the phenotypic description, and resulted in a molecular diagnosis. We subsequently tested the remainder of our diagnostic ES cohort, including over 30 clinically and genetically heterogeneous disorders. Optimized manual curation yielded 140 samples with a likely aberrant STR length. Validations confirmed 70/140 (50%) aberrant expansion alleles, of which 48 were in the pathogenic range and 22 in the premutation range.

**Conclusions:** Our work provides guidance for the implementation of STR analysis in clinical ES. Our results show that systematic STR evaluation may increase diagnostic ES yield by 0.2%, and recommend to make STR evaluation a routine part of ES interpretation in genetic testing laboratories.

## Introduction

Short tandem repeats (STRs), also called microsatellites, constitute ∼3% of the human genome and are scattered throughout^1,2^. STRs vary in size but are commonly defined as tandemly repeated nucleotide motifs of 2-12 base pairs in length^3^. Expansions of a subset of STRs can cause approximately 30 different human genetic disorders. A large group of repeat expansions cause different forms of spinocerebellar ataxias, while others cause Huntington disease (OMIM 143100), fragile X syndrome (OMIM 300624), or myotonic dystrophies (e.g. OMIM 160900 and OMIM 602668)^4^.

Next-generation sequencing (NGS) has proven to be of great diagnostic value in clinical practice^5,6^. Over the last years, several different tools for genome-wide genotyping of STRs from short read sequencing data, and mainly genome sequencing data, were developed^7-13^. Despite the extensive application of exome sequencing (ES) in routine diagnostic genetic testing and the many STR detection studies being published, STRs were not routinely identified from these data, and large-scale assessments of the diagnostic potential from detecting STRs from ES data have not yet been performed. In this study, we assess the clinical utility of detecting STR expansions in ES data for patients with (suspected) rare genetic disorders based on a cohort of 38,095 clinical exomes. We provide guidance for the implementation of a STR detection workflow in routine diagnostic clinical ES analysis with minimal additional analytic burden, and we show that it increases the ES diagnostic yield in our cohort of 38,095 clinical exomes.

## Material and Methods

### Tool selection on validation cohort

Based on the ability to call STRs from short read NGS data, we initially considered three different STR detection tools, STRetch^8^, ExpansionHunter^9^, and GangSTR^7^. We evaluated the performance using a validation cohort of 11 patients with known pathogenic repeat expansion allele(s) (**Supplementary Table 1**). These expansion alleles were previously identified using conventional genetic testing by fragment length analysis of PCR or repeat-primed PCR (RP-PCR) fragments. The three tools were locally installed using their GitHub installation page and were run according to the developer’s instructions.

### Selection of STR sites

STR loci of interest for this study were selected in three steps. First, we selected 23 well-described disease-causing STR loci from literature (**Supplementary Table 2**). Subsequently, we discarded *HTT* and *JPH3*, causing Huntington disease, a well-recognizable clinical phenotype, unlikely present in our cohorts, and thus preventing the risk on incidental findings. Finally, after running ExpansionHunter on the validation cohort, we removed the loci without sequencing coverage (n=5, **Supplementary Table 2**).

### Samples

In this study, two cohorts were analyzed, using ES data obtained as part of routine diagnostic work-up of patients with rare diseases suspected to have genetic origin (https://gdnm.nl/ for an overview of all 30 clinically and genetically heterogeneous disorders included). This study was approved by the institutional review board ‘Commissie Mensgebonden Onderzoek Regio Arnhem-Nijmegen’ under number 2011/188. All DNA samples were sequenced on an Illumina HiSeq 2000 instrument in combination with Agilent version 4 enrichment kit, or on Illumina HiSeq 4000 combined with an Agilent version 5 enrichment kit. More than 95% of samples had at least 75x median coverage across the enrichment kit targets. To optimize our STR detection approach, we used a de-identified ES cohort of 2,867 patients presenting with a movement disorder of suspected genetic origin, further referred to as the movement disorders cohort. For this cohort we only looked at STRs in genes associated with movement disorders (**Supplementary Table 3**). Subsequently, we detected STRs from our entire ES cohort of 38,095 anonymized samples, for which we used all selected STR loci of interest (**Supplementary Table 2**). The latter cohort also included the movement disorders cohort, but identical calls between the two cohorts were discarded to prevent double counting. In addition, this cohort contained both patients as well as unaffected individuals. We analyzed ES data as previously reported^15^.

### Analysis of the cohorts

Genotypes of each expansion locus were then compared to the locus specific expansion thresholds, which were set according to existing literature (**Supplementary Table 2**). Genotypes were classified in three different groups: normal range, grey zone range, and pathogenic range. The grey zone is defined as either uncertain whether there is an actual expansion, whether they are pathogenic, or whether there is incomplete penetrance.

### Confirmation of STRs

Alignments of the calls exceeding the grey zone threshold were manually curated for sequencing coverage, read mapping quality, and taking into account the specifics of the particular repeat structure, using the GraphAlignmentViewer^16^. All likely aberrant expansion alleles were tested using PCR and fragment length analysis using GeneScan^17^. When this only detected one normal allele size or indicated a pathogenic expansion allele, we also performed repeat-primed (RP-PCR) to confirm the GeneScan result or to make sure we did not miss a larger allele due to allelic dropout of normal PCR. RP-PCR for the *DMPK* and *CNBP* genes was performed as described^17^. For other loci, primer binding sites and test conditions are available upon request.

### Genetic diagnosis of patients

For the confirmed expansion alleles in the movement disorders cohort, the short clinical descriptions of the patients were compared to the OMIM phenotype of the concomitant disorders by a trained clinical laboratory geneticist.

## Results

ExpansionHunter demonstrated the highest sensitivity by detecting 10 (91%) of 11 known pathogenic repeat expansion alleles compared to GangSTR (73%) and STRetch (18%) (**Supplementary Figure 1; Supplementary Table 4**). We decided to use ExpansionHunter in our study, based on its higher sensitivity and specificity combination for the detection of aberrant STR lengths in our validation cohort.

### Systematic analysis of STRs in movement disorders cohort

To optimize our approach for detecting aberrant expansion alleles from ES data using ExpansionHunter in a diagnostic setting, we first applied the tool to the 2,867 patients in the movement disorders cohort, which yielded 87 aberrant STR alleles in 84 (2.9% of 2,867) different patients (**Figure 1a and 1c**; **Supplementary Table 5**). Manual curation allowed us to discard 51 expansion alleles, leaving 36 alleles with a likely aberrant STR length (**Figure 1c**). From the 36 samples with a likely repeat expansion, validation by PCR and/or repeat-primed PCR (RP-PCR) technologies confirmed the presence of aberrant expansion allele(s) in 11 (31% validation rate) (**Figure 1c and 1d**; **Supplementary Table 5 and 6**). For the 51 alleles that were discarded after manual inspection, we also performed validations to exclude false negative calls, but all of these samples showed repeat expansions within the normal range based on PCR and fragment length analysis.

**Figure 1.**
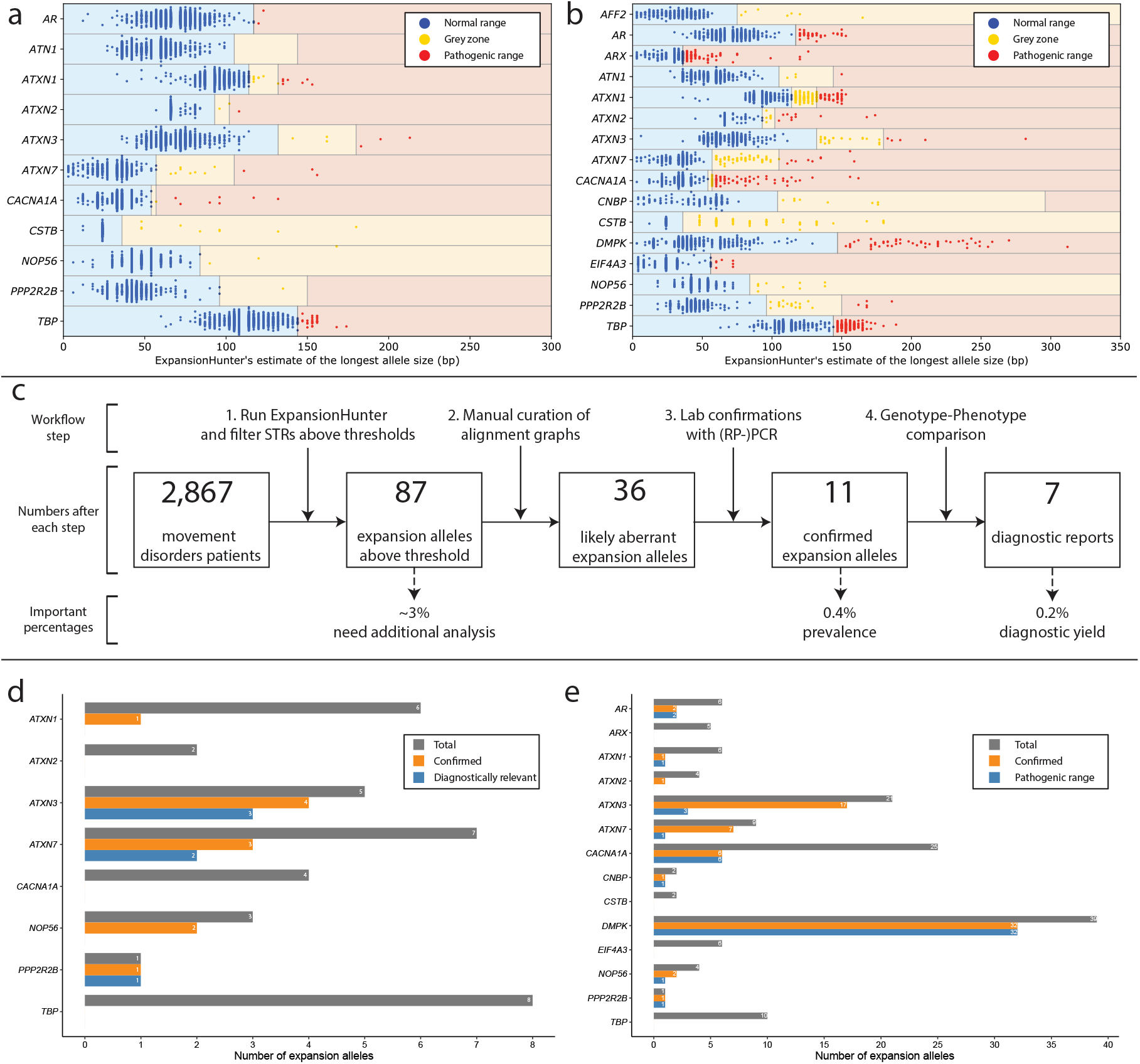
Results of STR size detection in two cohorts of ES samples and confirmations of alleles which exceeded the thresholds. **a)** Allele sizes for 2,867 ES samples of patients with movement disorders. Only movement disorder associated genes were analyzed. All samples in the movement disorders cohort with a STR length above the thresholds are listed in Supplementary Table 5. **b)** Allele sizes for all loci of interest for the full cohort of 38,095 ES samples. All samples in the full cohort with a STR length above the thresholds are listed in Supplementary Table 7. **c)** The workflow that we used to analyze the ES data of the 2,867 patients presenting with a movement disorder. The different steps are projected in the top part. The numbers of STR expansion alleles that were left after each step are presented in the middle and in the bottom part three concluding percentages are displayed. **d)** Validation rate per gene for the movement disorders cohort. **e)** Validation rate per gene for all 38,095 clinical ES samples.

Because of the 25 false positives and 67% specificity of the manual curation of the alignment graphs, we reanalyzed the graphs of the false positive calls and found that these graphs showed lower quality sequence flanking the STRs, more misaligned bases, and lower read depth (**Supplementary Figure 2**). Therefore, in order to optimize our workflow, we decided to improve the manual curation process of the STRs called by ExpansionHunter in the remainder of the full cohort.

### Aberrant STR calls in remainder of full cohort

Using the optimized approach for aberrant STR length detection, we systematically analyzed our full diagnostic ES cohort (**Figure 1b**). Among the 38,095 ES samples, we identified 1,091 aberrant expansion alleles in 1,080 different samples (2.8% of 38,095). Applying our stricter manual curation yielded 40 likely aberrant expansion alleles in 140 different samples (0.4% of 38,095) (**Supplementary Table 7 and 8**). Validation by PCR and/or RP-PCR confirmed the presence of 70 aberrant expansion alleles (50%) (**Figure 1e**). For the 70 confirmed expansion alleles, 48 were above the pathogenic repeat threshold, while 22 were in the grey/premutation zone. Notably, the application of our improved manual curation, significantly increased the validation rate for the likely aberrant expansion alleles compared to those achieved for the movement disorders cohort (*P*=0.041, two-sided Fisher’s exact test) (**Supplementary Table 6 and 8)**.

### Diagnostic implications of novel STR events

For 11 of the 2,867 samples from the movement disorders cohort, an aberrant expansion allele was detected by ExpansionHunter and confirmed by PCR and RP-PCR (**Figure 1c**). Of these, one expansion allele was in *ATXN1*, four in *ATXN3*, three in *ATXN7*, two in *NOP56* and one in *PPP2R2B* (**Figure 1d**), causing spinocerebellar ataxia 1, 3, 7, 36 and 12 respectively (OMIM 164400, OMIM 109150, OMIM 164500, OMIM 614153, OMIM 604326). Given the diverse genetic and clinical composition of our cohort, we evaluated whether the genetic findings correspond with the phenotype of the patient. For seven of the eleven samples the genotype was compatible with the phenotypic description and led to a genetic diagnosis for these patients (**Figure 1c; Figure 2**; **Supplementary Table 9**). For the other four confirmed expansion alleles, the genotype was not compatible with the clinical phenotype description. Based on this we estimate the additional diagnostic yield in this cohort of prescreened movement disorder patients to be at least 0.2% (7/2867).

**Figure 2.**
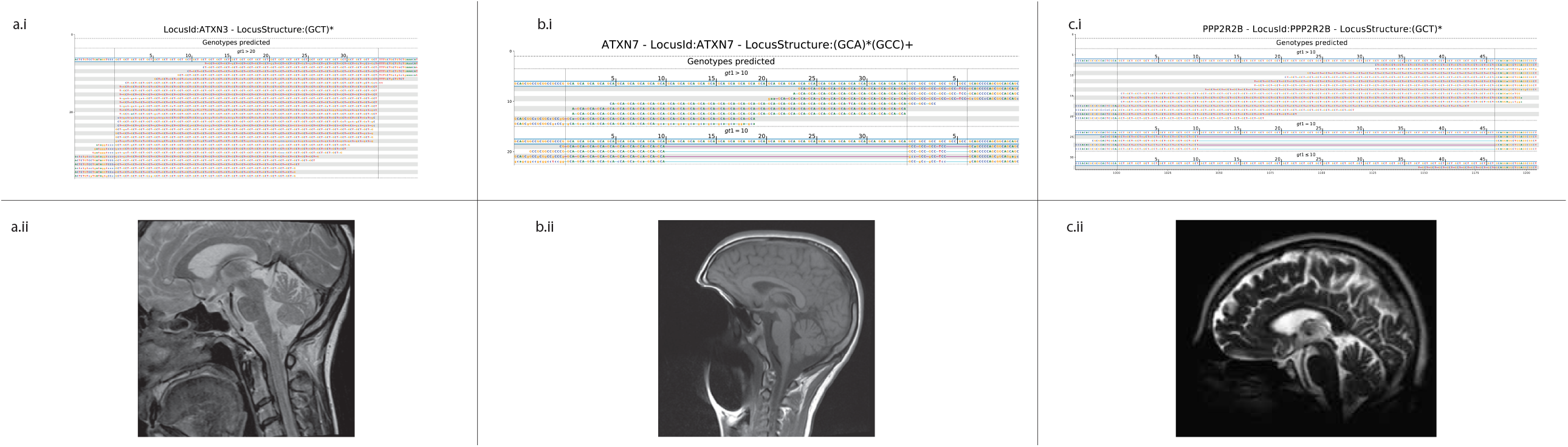
Three examples of clinical compatibility between genotype and the phenotypic description of two patients from the movement disorders cohort that received a novel genetic diagnosis based on the STR detection workflow, and one validation sample to compare to. For these three patients, the ExpansionHunter findings resulted in a novel genetic diagnosis or confirmed the previously detected expansion allele, showing the importance of routine diagnostic STR detection from ES data. For these patients a family tree is available upon request. **a.i)** ExpansionHunter alignment graph depicting the detected STR expansion in *ATXN3* in this patient.. **a.ii)** The sagittal T2 MRI brain scan showing atrophy of the cerebellar vermis. **b.i)** ExpansionHunter alignment graph for the second patient showing the repeat expansion in *ATXN7* for one of the validation samples. Running ExpansionHunter on this sample confirmed the presence of the previously identified expansion allele in *ATXN7* in this sample. **b.ii)** The sagittal T1 MRI brain scan showing absence of clear cerebellar vermian atrophy at the age of onset of the disorder for the index, despite the compatibility between the genotype and phenotypic description. **c.i)** ExpansionHunter alignment showing the repeat expansion in *PPP2R2B*. **c.ii)** The sagittal T2 MRI brain scan shows mild cerebellar but also cerebral cortical atrophy.

For the 70 confirmed expansions in the full cohort, the genotype could not be compared with the phenotypic description, since the samples in this cohort were anonymized to prevent the detection of incidental findings.

## Discussion

In this study, we report a systematic analysis of STRs in a large clinical ES dataset.

Notwithstanding the good diagnostic results we obtained with ExpansionHunter, we also show that adding a manual curation step to the workflow can be of great importance for filtering out false positive ExpansionHunter calls. In the future, the implementation of a robust quality control score for STRs may make this curation obsolete, which would lower the risk of interpersonal interpretation and increase the diagnostic robustness. However, the number of ExpansionHunter calls that were manually curated was only 87 for the movement disorders cohort (1 in 33 samples) and 1,091 for the full cohort (1 in 35 samples) and this means that the burden of additional analysis in routine diagnostics can be minimized, with only ∼3% of the samples requiring dedicated follow-up. Therefore, this should not hamper diagnostic implementation. Complementary experimental validations will however remain necessary to confirm detected expansion alleles.

The prevalence of aberrant expansion alleles was higher in our movement disorders cohort compared to our diagnostic ES cohort (0.4% vs 0.2% respectively), likely due to the fact that the ES cohort also contained samples of unaffected individuals (e.g. parents). In addition, it is known that STRs play a major role in the disease etiologies of movement disorders (mainly spinocerebellar ataxias), and therefore this cohort may be enriched for STR expansions^3^. Still the diagnostic yield for the movement disorders cohort was only 0.2% (7/2,867), which is likely due to the clinical pre-screening of this group of patients for repeat expansions.

Seven cases of the movement disorders cohort with a confirmed STR expansion received a genetic diagnostic report which will help them provide insight into the prognosis of the disorder and help patients and their relatives understand the cause of the disorder that is segregating in their family. For the other four confirmed expansion alleles, the genotype was not compatible with the clinical phenotype description. This is partly due to the thresholds we used for filtering the ExpansionHunter calls. Patients with a small or premutation-sized repeat expansion allele may not present with characteristic clinical features of the specific disorder yet, such as for example repeat expansions in the *DMPK* gene, causing myotonic dystrophy type 1. This disorder shows extreme anticipation with varying clinical features for small premutation (36-49 repeats; no disease), medium (50-150 repeats, mild and or late onset) or large allele sizes (>150 repeats; spectrum from late onset to congenital disease)^17^. In agreement, the detected pathogenic *DMPK* alleles in our anonymized cohort were all medium. Genetic diagnostic laboratories may need to consider carefully about how to manage such findings.

## Data Availability

All available data is included within the manuscript.

## Acknowledgments

We thank Michael Eberle and Egor Dolzhenko for kindly providing the Python code for the swimlane plots. This project was financially supported by an Aspasia grant of the Dutch Research Council (015.014.066 to LELMV), a VIDI grant (917-17-353 to CG) and the NWO X-omics project (184.034.019 to CG). The aims of this study contribute to the Solve-RD project (to CG and LELMV) which has received funding from the European Union’s Horizon 2020 research and innovation programme (No 779257).

## Code availability

ExpansionHunter script is available at: https://github.com/Illumina/ExpansionHunter

GangSTR script is available at: https://github.com/gymreklab/GangSTR

STRetch script is available at: https://github.com/Oshlack/STRetch

## Author Contribution Statement

**Bart P.G.H. van der Sanden:** Methodology, Project administration, Writing - original draft, Writing – review & editing. **Jordi Corominas:** Methodology, Software, Formal analysis, Investigation, Data curation. **Michelle de Groot:** Methodology, Software, Formal analysis, Investigation, Data curation. **Maartje Pennings:** Validation, Formal analysis. **Rowdy P.P. Meijer:** Validation, Formal analysis. **Nienke Verbeek:** Resources, Visualization. **Bart van de Warrenburg:** Resources, Visualization. **Meyke Schouten:** Resources, Visualization. **Helger G. Yntema:** Writing – review & editing. **Lisenka E.L.M Vissers:** Writing – review & editing, Funding acquisition. **Erik-Jan Kamsteeg:** Conceptualization, Methodology, Project administration, Writing – original draft, Writing – review & editing, Supervision. **Christian Gilissen:** Conceptualization, Methodology, Project administration, Writing – original draft, Writing – review & editing, Supervision, Funding acquisition.

## Ethics declarations

### Disclosure

The authors declare no conflict of interest.

## Notes

### Competing Interest Statement

The authors have declared no competing interest.

### Author Declarations

Patient cohorts provided relevant informed consent or where anonymized for this study. This study was approved by the institutional review board Commissie Mensgebonden Onderzoek Regio Arnhem-Nijmegen under number 2011/188.

