## Supplementary Text for "Systematic analysis of short tandem repeats in 38,095 exomes provides an additional diagnostic yield"

**Supplementary Legends**

**Supplementary Figure 1.** ExpansionHunter visualization of the repeat expansion in the gene *ATXN2* (normal allele of 22 repeats, pathogenic allele of 39 repeats) in one sample that was among the 11 validation samples, but was not detected by ExpansionHunter, due to low sequence coverage. Repeat expansion locus consists of the left and right flanking sequence with the repeated GCT expansion motif in the middle (i.e. in between vertical lines). Relevant reads are extracted from the sequencing data and are aligned to the locus structure. Subsequently, genotypes are calculated.

**Supplementary Figure 2.** Examples of visual output by ExpansionHunter that was manually inspected. **a)** Alignment graph of a sample with a confirmed repeat expansion at the *ATXN7* locus. Flanking sequence is of good quality and well aligned. Also, the repeat motif is consistent and not interrupted. Finally, there are very little mismatched bases in the aligned repeat reads. **b)** Alignment graph of a sample that was filtered out during manual curation. Flanking sequence is absent or not well aligned. In addition, the repeat reads contain multiple mismatches and interruptions as well as different repeat motifs. Finally, read depth for longest allele size is very low and second largest allele size is below the threshold of 19.

**Supplementary Tables**

**Supplementary Table 1.** Overview of eleven samples with known repeat expansions used in the validation cohort.

| **Sample** | **Gene name** | **Disease symbol** | **Expansion** | **Identified by** | **Sequencing instrument** | **Enrichment kit** | **Median coverage** |
| --- | --- | --- | --- | --- | --- | --- | --- |
| S7652 | *ATXN1* | SCA1 | 41 | EH, GangSTR | HiSeq4000 | Agilent V5 | 118 |
| S26152 | *ATXN2* | SCA2 | 39 | GangSTR | HiSeq4000 | Agilent V5 | 107 |
| S37923 | *ATXN3* | SCA3 | 56 | EH, STRetch | HiSeq4000 | Agilent V5 | 110 |
| S31471 | *ATXN7* | SCA7 | 58 | EH, GangSTR | HiSeq2000 | Agilent V4 | 115 |
| S8656 | *ATXN7* | SCA7 | 58 | EH, STRetch | HiSeq2000 | Agilent V4 | 98 |
| S28976 | *CACNA1A* | SCA6 | 22 | EH, GangSTR | HiSeq4000 | Agilent V5 | 102 |
| S23830 | *CACNA1A* | SCA6 | 23 | EH, GangSTR | HiSeq4000 | Agilent V5 | 118 |
| S34960 | *CACNA1A* | SCA6 | 23 | EH, GangSTR | HiSeq4000 | Agilent V5 | 114 |
| S13979 | *CACNA1A* | SCA6 | 23 | EH, GangSTR | HiSeq4000 | Agilent V5 | 122 |
| S15569 | *CACNA1A* | SCA6 | 22 | EH, GangSTR | HiSeq4000 | Agilent V5 | 105 |
| S25653 | *TBP* | SCA17 | 56 | EH | HiSeq4000 | Agilent V5 | 99 |

| **Gene name** | **Symbol** | **Inheritance** | **Reference** | **Genomic location** | **Location in gene** | **Repeat unit** | **Normal repeat size (repeat units)** | **Greyzone repeat size (repeat units)** | **Pathogenic repeat size (repeat units)** | **Threshold (repeat units)** |
| --- | --- | --- | --- | --- | --- | --- | --- | --- | --- | --- |
| *AFF2* | FRAXE | XL | [1] | chrX:147582151-147582211 | 5'UTR | GCC | 6-25 | 26-200 | >200 | 25 |
| *AR* | AIS | XL | [2] | chrX:66765158-66765227 | Exon | GCA | 17-26 | 27-39 | >39 | 39**^a^** |
| *ARX* | EIEE1 | XL | [3] | chrX:25031779-25031808 | Exon | CGC | 9-12 | - | >12 | 12 |
| *ATN1* | DRPLA | AD | [4] | chr12:7045879-7045936 | Exon | CAG | 6-35 | 36-48 | >48 | 35 |
| *ATXN1* | SCA1 | AD | [4] | chr6:16327864-16327954 | Exon | TGC | 6-38 | 39-44**^b^** | >44 | 38 |
| *ATXN10** | SCA10 | AD | [4] | chr22:46191234-46191304 | Intron | ATTCT | 8-32 | 33-850 | >850 | 32 |
| *ATXN2* | SCA2 | AD | [4] | chr12:112036753-112036822 | Exon | GCT | 14-31 | 32-34 | >34 | 31 |
| *ATXN3* | SCA3 | AD | [4] | chr14:92537353-92537386 | Exon | GCT | 11-44 | 45-60 | >60 | 44 |
| *ATXN7* | SCA7 | AD | [4] | chr3:63898360-63898390 | Exon | GCA | 4-19 | 20-35 | >35 | 19 |
| *C9orf72** | FTDALS1 | AD | [5] | chr9:27573526-27573544 | Intron | GGCCCC | 2-19 | 20-250 | >250 | 19 |
| *CACNA1A* | SCA6 | AD | [4] | chr19:13318672-13318711 | Exon | CTG | 4-18 | 19 | >19 | 18 |
| *CNBP* | DM2 | AD | [6] | chr3:128891419-128891575 | Intron | CAGG | <27 | 27-74 | >74**^c^** | 26 |
| *CSTB* | EPM1A | AR | [7] | chr21:45196324-45196360 | 5'UTR | CGCGGGGCGGGG | 2-3 | 4-60 | >60 | 3 |
| *DMPK* | DM1 | AD | [8] | chr19:46273462-46273522 | 3'UTR | CAG | 5-35 | 36-49 | >49 | 49**^a^** |
| *EIF4A3* | RCPS | AR | [9] | chr17:78120972-78120995 | 5'UTR | CGTG | 3-12 | 13 | >13 | 14**^a^** |
| *FMR1** | FXS | XL | [10] | chrX:146993568-146993628 | 5'UTR | CGG | 5-44 | 45-200 | >200 | 45 |
| *FXN** | FRDA | AR | [11] | chr9:71652202-71652220 | Intron | GAA | 5-33 | 34-65 | >65 | 33 |
| *NOP56* | SCA36 | AD | [12] | chr20:2633379-2633403 | Intron | GGCCTG | 3-14 | 15-670 | >670 | 14 |
| *PPP2R2B* | SCA12 | AD | [4] | chr5:146258290-146258320 | 5'UTR | GCT | 4-32 | 33-50 | >50 | 32 |
| *RFC1** | CANVAS | AR | [13] | chr4:39350044-39350099 | Intron | AAGGG | - | - | >400**^c,d^** | + |
| *TBP* | SCA17 | AD | [4] | chr6:170870994-170871105 | Exon | GCA | 25-42 | 43-48 | >48 | 48**^a^** |

**Supplementary Table 2.** List of medically relevant repeats, pathogenic ranges and repeat thresholds used as loci of interest in this study.

(a) the threshold was set for the pathogenic repeat size due to the large amount of false-positives when using the greyzone threshold; (-) unknown; (b) repeats of this size are pathogenic when pure, not interrupted by CAT units; (c) Smallest pathogenic repeat size detected in a patient; (b) the normal RFC1 repeat has units of AAAAG or AAAGG, no small repeats with the pathogenic AAGGG unit are known; (+) when detected; greyzone is defined as either uncertain whether these repeat sizes exist, whether they are pathogenic, or incomplete penetrance; (*) loci without sequencing coverage in ES data due to the gene component location of the locus in the gene; References refer to supplementary references.

**Supplementary Table 3.** Disorders with associated genes in which coding repeat expansions are medically relevant.

| **Gene panel (STR)** | **Gene names** |
| --- | --- |
| Movement disorders | *AR, ATN1, ATXN1, ATXN2, ATXN3, ATXN7, ATXN10, C90rf72, CACNA1A, CSTB, FXN, NOP56, PPP2R2B, RFC1, TBP* |
| Vision disorders | *ATXN7* |
| Epilepsy | *CSTB* |
| Hemostatic/thrombotic disorders | *AR* |
| Intellectual disability | *AFF2, ARX, DMPK, EIF4A3, FMR1* |
| Muscle disorders | *AR, CNBP, DMPK* |

**Supplementary Table 4.** Summary of the comparison of all allele sizes of the STRs in the genes: *ATXN1, ATXN2, ATXN3, ATXN7, CACNA1A* and *TBP.* Sensitivity is TP/(TP+FN). Specificity is TN/(TN+FP).

| **Tool** | **True positives (TP)** | **False positives (FP)** | **True negatives (TN)** | **False negatives (FN)** | **Total** | **Sensitivity** | **Specificity** |
| --- | --- | --- | --- | --- | --- | --- | --- |
| **GangSTR** | 8 | 0 | 55 | 3 | 66 | 73% | 100% |
| **ExpansionHunter** | 10 | 0 | 46 | 1 | 57 | 91% | 100% |
| **STRetch** | 2 | 0 | 55 | 9 | 66 | 18% | 100% |

**Supplementary Table 5.** Overview of identified STRs by ExpansionHunter in a cohort of 2,867 patients with movement disorders.

*External file: Table 5 - STR_patients_movementDisorders.xlsx*

**Supplementary Table 6.** GeneScan and RP-PCR validation results for 36 likely aberrant expansion alleles from 2,867 ES samples of patients with movement disorders. Confirmed means GeneScan and RP-PCR confirmed the presence of the expansion allele that was called by ExpansionHunter. Not confirmed means the expansion allele was not confirmed by GeneScan and RP-PCR.

| **Gene** | **Confirmed** | **Not Confirmed** | **Total** | **Percentage Confirmed** |
| --- | --- | --- | --- | --- |
| *ATXN1* | 1 | 5 | 6 | 17 |
| *ATXN2* | 0 | 2 | 2 | 0 |
| *ATXN3* | 4 | 1 | 5 | 80 |
| *ATXN7* | 3 | 4 | 7 | 43 |
| *CACNA1A* | 0 | 4 | 4 | 0 |
| *NOP56* | 2 | 1 | 3 | 67 |
| *PPP2R2B* | 1 | 0 | 1 | 100 |
| *TBP* | 0 | 8 | 8 | 0 |
| Total | 11 | 25 | 36 | 42 |

**Supplementary Table 7.** Overview of identified STRs by ExpansionHunter in entire exome sequencing cohort of 38,095 patients with movement disorders.

*External file: Supplementary Table 7 - STR_patients_wholecohort.xlsx*

**Supplementary Table 8.** GeneScan and RP-PCR validation results for 140 likely aberrant expansion alleles from 38,095 ES samples. Confirmed means GeneScan and RP-PCR confirmed the presence of the expansion allele that was called by ExpansionHunter. Not confirmed means the expansion allele was not confirmed by GeneScan and RP-PCR.

| **Gene** | **Confirmed** | **Not Confirmed** | **Total** | **Percentage Confirmed** |
| --- | --- | --- | --- | --- |
| *AR* | 2 | 4 | 6 | 33 |
| *ARX* | 0 | 5 | 5 | 0 |
| *ATXN1* | 1 | 5 | 6 | 17 |
| *ATXN2* | 1 | 3 | 4 | 25 |
| *ATXN3* | 17 | 4 | 21 | 81 |
| *ATXN7* | 7 | 2 | 9 | 78 |
| *CACNA1A* | 6 | 19 | 25 | 24 |
| *CNBP* | 1 | 1 | 2 | 50 |
| *CSTB* | 0 | 2 | 2 | 0 |
| *DMPK* | 32 | 7 | 39 | 82 |
| *EIF4A3* | 0 | 6 | 6 | 0 |
| *NOP56* | 2 | 2 | 4 | 50 |
| *PPP2R2B* | 1 | 1 | 1 | 50 |
| *TBP* | 0 | 10 | 10 | 0 |
| Total | 70 | 70 | 140 | 50 |

**Supplementary Table 9.** Diagnostic results for identified pathogenic repeat expansions from ES.

| **SampleID** | **Gene** | **Disease** | **Expansion size ExpansionHunter (repeat units)** | **Expansion size (RP)-PCR (repeat units)** | **Gene Panel** | **Clinical Diagnosis** | **Clinical compatibility** | **new** | **Comment** |
| --- | --- | --- | --- | --- | --- | --- | --- | --- | --- |
| S21020 | ATXN1 | SCA1 | 34/40 | 33/39 | mov.dis. | mother of child with mild facial dysmorphic features, low tone, tics | uncertain | y | With interruption |
| S3856 | ATXN3 | SCA3 | 20/65 | 23/65 | mov.dis. | sporadic CA | yes | y |  |
| S7285 | ATXN3 | SCA3 | 21/71 | 24/63 | mov.dis./HMSN | AD CA / neuropathy | yes | y |  |
| S20251 | ATXN3 | SCA3 | 24/61 | 27/53 | mov.dis. | CA / HSP | yes | n |  |
| S20755 | ATXN3 | SCA3 | 54 | 14/51 | mov.dis. | tremor / myocl. / dyst. | uncertain | y |  |
| S17831 | ATXN7 | SCA7 | 31 | 10/43 | mov.dis. | CA / HSP | yes | n |  |
| S34084 | ATXN7 | SCA7 | 11/52 | 12/66 | mov.dis. / vision disorders | CA / HSP / CRD /RD | yes | y |  |
| S28638 | ATXN7 | SCA7 | 24 | 10/24 | mov.dis. | father* of child with HSP and hyperventilation | uncertain | y |  |
| S913 | NOP56 | SCA36 | 7/20 | 5/15 | mov.dis. | muscle weakness in legs and intellectual disability | uncertain | y |  |
| S8867 | NOP56 | SCA36 | 7/15 | 5/14 | mov.dis. | CA | yes | y |  |
| S17070 | PPP2R2B | SCA12 | 45 | 9/54 | mov.dis. | progressive neurodegenerative disease, essential tremor | yes | y |  |

*paternal bias in expansion

myotonic dystrophy 1/2 (MD1/2), movement disorders (mov.dis.), hereditary motor- and sensory neuropathy (HMSN), autosomal dominant (AD), spinocerebellar ataxia (SCA), hereditary spastic paraplegia (HSP), myoclonia (myocl.), dystonia (dyst.), cone-rod dystrophy (CRD), retina dystrophy (RD). Column “New?” indicating whether diagnosis was new or previously known.
