## Supplementary figures and images for "Systematic analysis of short tandem repeats in 38,095 exomes provides an additional diagnostic yield"

### Supplementary Figure 1

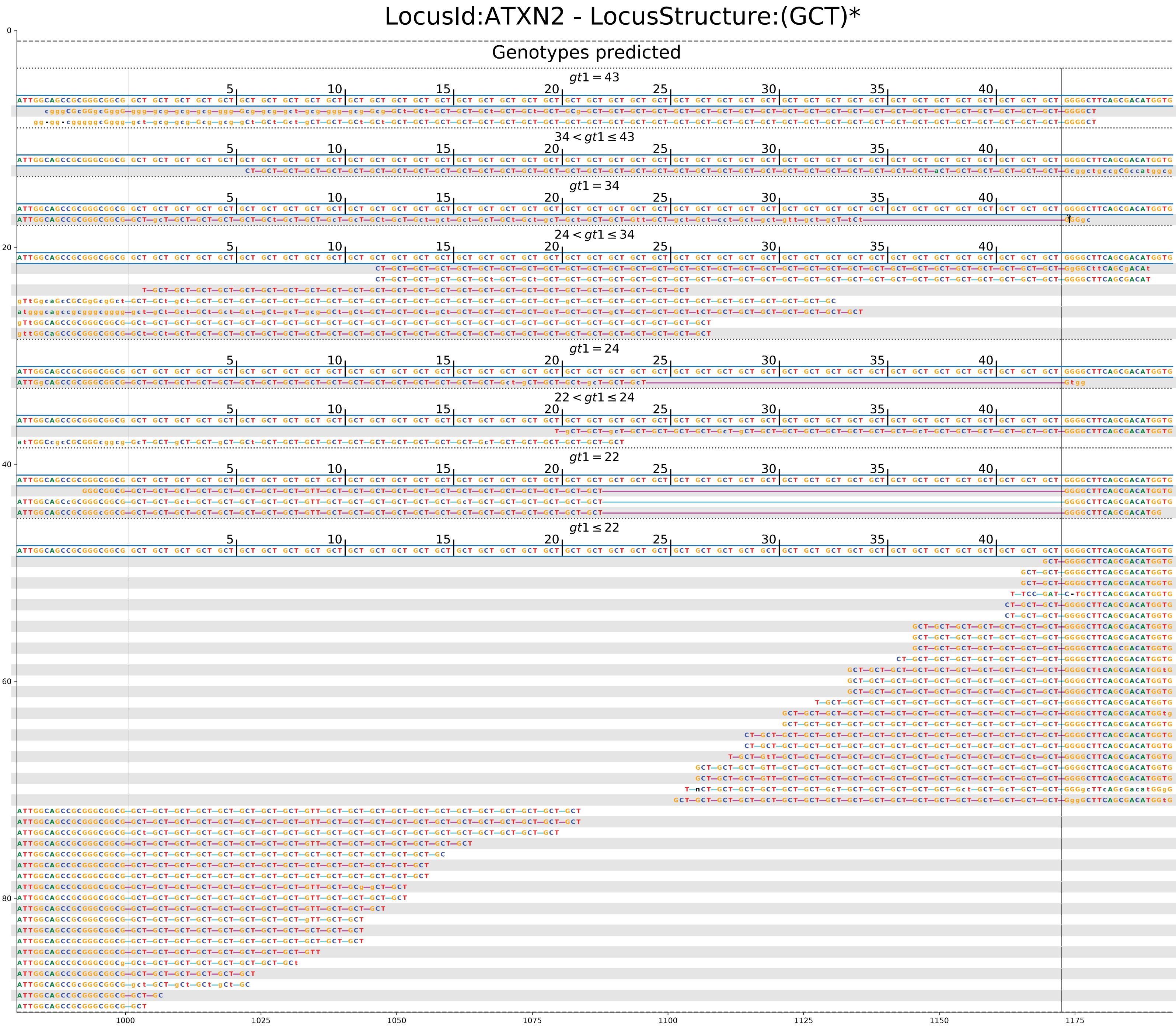

### Supplementary Figure 2

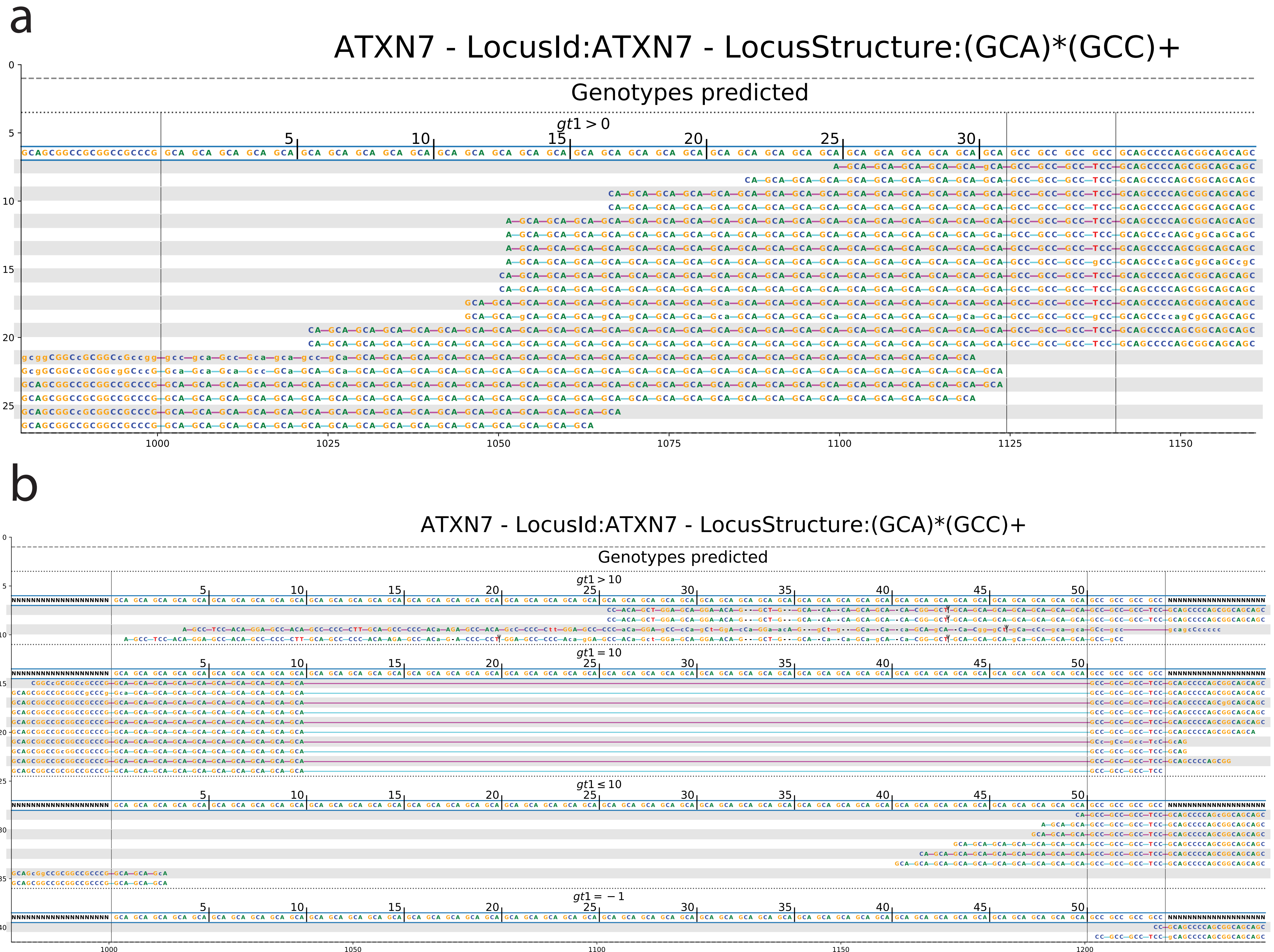
